# AI-Powered Triage of Suicidal Ideation in Adolescents: A Comparative Evaluation of Large Language Models Using Synthetic Clinical Vignettes

**DOI:** 10.1101/2025.08.05.25333046

**Authors:** Masab Mansoor

## Abstract

**Objective:** To evaluate the performance of leading Large Language Models (LLMs) in classifying suicide risk and generating clinically appropriate action plans for adolescent psychiatric cases presented through synthetic clinical vignettes.

**Methods:** We developed 40 synthetic clinical vignettes depicting adolescents with varying levels of suicide risk, structured according to established clinical formulation principles. A gold standard for risk level, based on the Columbia-Suicide Severity Rating Scale (C-SSRS) framework, and corresponding clinical actions was established for each vignette by a panel of two board-certified child and adolescent psychiatrists. Three LLMs (GPT-4o, Claude 3.5 Sonnet, Llama-3.1-70B) were prompted using a structured chain-of-thought methodology to classify risk and propose a detailed action plan. Performance was assessed using quantitative classification metrics (accuracy, precision, recall, F1-score) and qualitative thematic analysis of the generated action plans.

**Results:** Quantitative analysis of risk classification revealed variable performance. GPT-4o achieved the highest accuracy (82.5%), followed by Claude 3.5 Sonnet (75.0%) and Llama-3.1-70B (67.5%). F1-scores demonstrated challenges in correctly identifying higher-risk categories, particularly for nuanced presentations of intent. Qualitative thematic analysis of the action plans identified consistent adherence to basic safety protocols (e.g., recommending emergency evaluation for high-risk cases). However, significant and critical failures were pervasive, including the omission of crucial inquiries about access to lethal means, failure to incorporate protective factors into planning, and the generation of clinically inappropriate therapeutic reassurance in a triage context.

**Conclusions:** While LLMs demonstrate a nascent ability to process clinical information for suicide risk assessment, significant deficits in clinical reasoning and safety planning persist. Their performance on idealized synthetic data suggests these models are not yet suitable for autonomous clinical decision-making. These findings underscore the imperative for rigorous, clinically-grounded evaluation frameworks and the development of human-in-the-loop systems to ensure patient safety in any future deployment.

**Key Messages:** *What is already known on this topic:* Suicide is a leading cause of death in adolescents, yet current clinical risk assessment tools and subjective judgments have limited predictive accuracy and are difficult to scale in the face of rising demand and workforce shortages.

*What this study adds:* This study provides a direct comparative evaluation of multiple state-of-the-art Large Language Models on a standardized adolescent suicide risk triage task, using a synthetic data methodology that allows for controlled assessment of both classification accuracy and the clinical appropriateness of generated action plans.

*How this study might affect research, practice or policy:* The findings highlight the potential of LLMs as adjunctive tools in non-specialist settings but also reveal critical safety and reliability gaps that must be addressed through further research, the development of ethical guidelines, and regulatory oversight before any clinical implementation can be considered.

## Introduction

### The Escalating Crisis of Adolescent Suicide

Suicide represents a profound and escalating public health crisis, particularly among young people. Globally, it is the third leading cause of death for adolescents aged 15–29.^1^ In the United States, the statistics are similarly dire, with suicide ranking as the second leading cause of death for individuals aged 10–34.^2^ Data from the Centers for Disease Control and Prevention (CDC) indicate that suicide deaths among 10- to 24-year-olds increased by 62% from 2007 to 2021.^4^ This trend is paralleled by a rise in suicidal ideation and attempts; in 2023, one in five high school students seriously considered attempting suicide ^4^, and 9% made at least one attempt in the past year.^2^

This crisis does not affect all populations equally. Disparities are stark, particularly for LGBTQ+ youth, who report significantly higher rates of suicidal ideation (39%) and attempts (12%) compared to their cisgender and heterosexual peers.^5^ Youth of color, including Native/Indigenous (24% attempted) and Black/African American (14% attempted) youth, also face disproportionately high rates.^5^ This surge in suicidality occurs alongside a broader decline in adolescent mental health, with nearly 1 in 5 teens experiencing a major depressive episode in the past year.^6^ The connection is clear: untreated or undertreated mental illness is a primary driver of suicide risk.^1^

### Deficiencies in Current Risk Assessment Paradigms

Despite the urgency of this crisis, the “standard of care” for suicide risk assessment is fraught with limitations. The process heavily relies on a clinician’s subjective judgment, a method with notoriously poor predictive accuracy for a low-base-rate event like suicide.^7^ It is widely acknowledged that predicting whether a specific individual will die by suicide is impossible; the clinical goal is assessment of risk to guide intervention, not a definitive forecast.^7^

Validated screening instruments, such as the Ask Suicide-Screening Questions (ASQ) and the Columbia-Suicide Severity Rating Scale (C-SSRS), have been widely adopted to standardize this process.^9^ However, these tools are not panaceas. Systematic reviews have shown that available assessment tools do not reliably predict future self-harm or suicide and often perform well on either sensitivity or specificity, but rarely both.^8^ This trade-off has significant consequences: low sensitivity leads to missed cases (false negatives), compromising patient safety, while low specificity leads to over-identification (false positives), which can strain already limited mental health resources.^8^ Furthermore, research has demonstrated that single-item screens or the use of depression screening as a proxy for suicide risk is insufficient, as these methods can miss a substantial portion of at-risk individuals who may not present with classic depressive symptoms.^13^

### The Emergence of AI as a Potential Adjunct in Psychiatric Triage

The convergence of the adolescent mental health crisis and the limitations of current assessment methods creates a critical need for innovative solutions. Artificial intelligence (AI), and specifically Large Language Models (LLMs), have emerged as powerful technologies capable of processing vast amounts of unstructured text data to identify patterns and support clinical decision-making.^15^ The potential applications in mental health are compelling. AI could help address workforce shortages by automating administrative tasks, freeing clinicians to focus on patient care.^18^ LLMs, trained on massive text corpora, can analyze clinical notes, patient communications, and social media data to detect signs of distress or risk.^20^ Early studies have shown promise in using LLMs to detect suicidal ideation from text, suggesting a potential role in risk prediction and triage.^22^

### Study Rationale, Objectives, and Hypotheses

This technological promise presents a critical paradox of scalability versus safety. While LLMs offer an unprecedented ability to scale mental health screening, this also creates the risk of deploying flawed or biased systems at scale, potentially causing widespread harm.^25^ There remains a significant gap in the literature regarding the rigorous, comparative evaluation of modern LLMs for the specific, high-stakes task of adolescent suicide risk triage. Critically, evaluation must extend beyond simple classification accuracy to assess the clinical appropriateness and safety of the actions these models recommend.

This study was designed to address this gap. The primary objective was to evaluate and compare the performance of three prominent LLMs (GPT-4o, Claude 3.5 Sonnet, and Llama-3.1-70B) against a clinical gold standard in two domains: (1) classifying the level of suicide risk, and (2) generating clinically appropriate and safe action plans. The evaluation was conducted using a set of purpose-built synthetic adolescent psychiatric vignettes, a methodology that allows for controlled and systematic testing without the ethical and privacy constraints of using real patient data.

We formulated three hypotheses: (1) LLM performance in risk classification would vary, with the more advanced proprietary models (GPT-4o, Claude 3.5 Sonnet) outperforming the open-weight model (Llama-3.1-70B); (2) all models would demonstrate moderate-to-high accuracy in classifying cases at the extremes of risk (i.e., no risk versus very high risk) but would struggle with more nuanced, intermediate-risk presentations; and (3) a qualitative analysis of the LLM-generated action plans would reveal significant deficiencies in clinical reasoning and safety planning when compared to the gold standard established by expert clinicians.

## Methods

### Study Design

This study employed a comparative, in-silico evaluation design. This approach involves testing computational models on standardized, synthetic data to assess performance in a controlled environment.^27^ This design was selected to enable the systematic manipulation of clinical variables within the vignettes and to allow for a direct comparison of LLM outputs against a predefined gold standard, thereby avoiding the ethical complexities and regulatory requirements associated with using real patient data.^28^ A study flow diagram, modeled after the CONSORT statement, illustrates the process from vignette development to final data analysis (Figure 1).^28^

**Figure 1.**
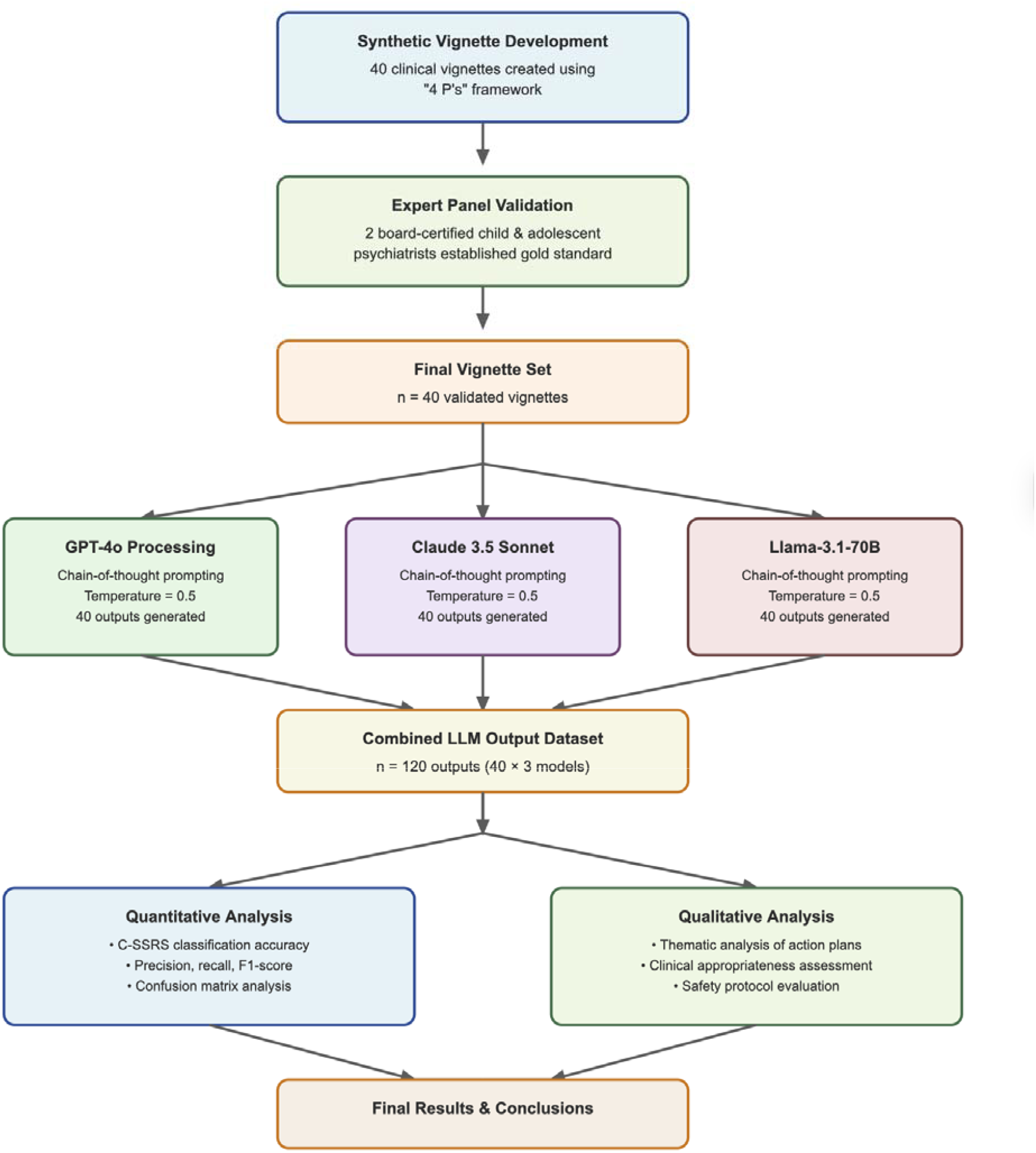
Study Flow Diagram. Flow diagram showing the progression from vignette creation, LLM processing, to data analysis.

### Synthetic Vignette Development and Validation

A set of 40 synthetic clinical vignettes was developed, adhering to established best practices for vignette methodology in health research.^29^ Vignettes are a recognized and cost-effective method for assessing clinical decision-making and exploring variations in practice.^30^ Each vignette was structured using the “4 P’s” of clinical formulation to ensure clinical depth and realism: **P**redisposing factors (e.g., family history of suicide, chronic illness), **P**recipitating factors (e.g., academic pressure, recent interpersonal conflict), **P**erpetuating factors (e.g., ongoing bullying, substance use, social isolation), and **P**rotective factors (e.g., strong family support, engagement in extracurricular activities, future-oriented thinking).^32^

To ensure a comprehensive test set, variables within the vignettes were systematically controlled and manipulated.^33^ Controlled variables included the patient’s age range (13–17 years), the clinical setting (e.g., emergency department, outpatient pediatric clinic, school counselor’s office), and the narrative perspective (e.g., a third-person psychiatric consult request, a first-person account from a concerned parent). Key experimentally manipulated variables included the severity of suicidal ideation (from passive wishes to be dead to active ideation with a specific plan and intent), history of prior suicide attempts, presence and nature of non-suicidal self-injury (NSSI), and the co-occurrence of significant risk factors such as substance use disorders, psychosis, impulsivity, and access to lethal means.^7^

The full set of vignettes underwent a content validation process, wherein a panel of two board-certified child and adolescent psychiatrists independently reviewed each case for clinical plausibility, realism, clarity, and internal consistency.^30^ Any disagreements were resolved through consensus discussion. The distribution of key clinical characteristics across the final set of 40 vignettes is detailed in Table 1.

**Table 1.** Characteristics of the 40 Synthetic Clinical Vignettes.

| Characteristic | Category | Number of Vignettes (%) |
| --- | --- | --- |
| **C-SSRS Risk Level (Gold Standard)** | 1: Wish to be Dead | 5 (12.5%) |
|  | 2: Non-Specific Active Thoughts | 5 (12.5%) |
|  | 3: Active Ideation with Method | 10 (25.0%) |
|  | 4: Active Ideation with Intent | 10 (25.0%) |
|  | 5: Active Ideation with Plan & Intent | 5 (12.5%) |
|  | Actual Attempt (Recent) | 5 (12.5%) |
| **Key Risk Factors** | History of Prior Attempt | 12 (30.0%) |
|  | Presence of NSSI | 15 (37.5%) |
|  | Substance Use Mentioned | 14 (35.0%) |
|  | Psychotic Features Mentioned | 6 (15.0%) |
|  | Explicit Access to Means | 8 (20.0%) |
| **Key Protective Factors** | Strong Social/Family Support | 18 (45.0%) |
|  | Engaged in Activities/Hobbies | 20 (50.0%) |
|  | Future Orientation | 10 (25.0%) |

### Gold Standard Definition

For each of the 40 vignettes, the expert clinical panel established a two-part “gold standard.” First, a definitive risk classification was assigned using the hierarchical structure of the Columbia-Suicide Severity Rating Scale (C-SSRS).^11^ This scale provides clear, operationalized definitions for different levels of suicidal ideation and behavior, ranging from a passive wish to be dead to an active attempt. Second, the panel formulated a corresponding gold-standard clinical action plan for each vignette. These plans were based on established clinical practice guidelines from the American Psychiatric Association (APA) and the framework of the Suicide Assessment Five-Step Evaluation and Triage (SAFE-T) Pocket Card.^7^ The action plans were stratified by risk level and specified necessary interventions, such as ensuring patient safety (e.g., initiating 1:1 observation), determining the level of care (e.g., recommending immediate psychiatric hospitalization), and providing specific treatment recommendations (e.g., conducting means restriction counseling with the family).

### Large Language Model Selection and Configuration

Three state-of-the-art LLMs were selected to represent the current landscape of AI technology: GPT-4o (a leading proprietary model from OpenAI), Claude 3.5 Sonnet (a proprietary model from Anthropic noted for its advanced reasoning capabilities), and Llama-3.1-70B (a powerful open-weight model from Meta).^37^ This selection allows for a comparison across different model architectures and training philosophies.

A structured, multi-step prompt was engineered to guide the LLMs’ analytical process, moving beyond simple classification to elicit a form of transparent reasoning.^40^ This “chain-of-thought” (CoT) prompting technique is designed to break down a complex task into intermediate steps, improving performance and providing insight into the model’s decision-making process.^41^ The prompt given to each LLM for every vignette consisted of four parts:

1. **Role Assignment** “You are an expert child and adolescent psychiatrist specializing in suicide risk assessment.” ^42^
2. **Context and Task Definition** “You will be provided with a clinical vignette describing an adolescent patient. Your task is to conduct a comprehensive suicide risk assessment and formulate a detailed, step-by-step clinical action plan.” ^43^
3. **Structured Reasoning Steps (CoT)** “To complete this task, you must proceed in the following order. First, analyze the vignette and explicitly list all identifiable risk factors and protective factors. Second, based on your analysis, classify the patient’s current suicide risk level using the specific categories of the Columbia-Suicide Severity Rating Scale (C-SSRS). Third, based on your risk classification, provide a clear, actionable, step-by-step clinical action plan.” ^40^
4. **Output Formatting** “Structure your final response with the following distinct headings: ‘Identified Risk and Protective Factors’, ‘C-SSRS Risk Classification’, and ‘Clinical Action Plan’.”

Each of the 40 vignettes was submitted as input to each of the three LLMs. To ensure reproducibility while allowing for coherent text generation, a consistent temperature setting of was used for all model queries.^44^ All prompts and the corresponding full-text outputs were logged for subsequent analysis.

### Ethical Considerations

This study exclusively utilized synthetically generated data that contained no personal or identifiable information of any real individuals. As such, the research did not involve human participants, and according to institutional and international guidelines, did not require review and approval from an ethics committee or institutional review board.

### Outcome Measures and Analysis

The evaluation of the LLM outputs was conducted through a two-pronged approach. First, a **quantitative analysis** was performed on the risk classification component. The C-SSRS classification generated by each LLM was compared against the expert-defined gold standard for each vignette. Standard performance metrics for multi-class classification were calculated: overall accuracy, and class-specific precision, recall, and F1-score.^39^ A confusion matrix was also generated for each model to provide a visual representation of misclassification patterns. Second, a **qualitative analysis** was conducted on the LLM-generated clinical action plans. This analysis is critical, as the appropriateness of the recommended actions is arguably more clinically significant than the classification label itself. A rigorous thematic analysis was performed on the 120 generated action plans (40 vignettes x 3 LLMs).^46^ Two independent researchers (a clinical psychologist and a psychiatric resident), blind to the LLM that generated each plan, coded the content using a hybrid deductive-inductive approach. The deductive coding framework was based on established safety guidelines ^7^ and evaluated the plans across three core domains: (1) **Safety Interventions** (e.g., presence of recommendations for immediate safety measures like removing access to means or ensuring constant observation); (2) **Assessment Comprehensiveness** (e.g., inclusion of steps like involving family/caregivers, conducting a full psychiatric evaluation); and (3) **Disposition Appropriateness** (e.g., proportionality of the recommended level of care to the identified risk). An inductive approach was used concurrently to identify emergent themes not captured by the deductive framework, such as the generation of inappropriate therapeutic language or model “hallucinations”.^37^ Inter-rater reliability (Cohen’s Kappa) was calculated for the deductive codes, with any discrepancies resolved through discussion with a third senior researcher (Author B).

## Results

### Study Flow

The study process is summarized in Figure 1. A total of 40 unique synthetic vignettes were created and validated. Each vignette was processed by the three selected LLMs, resulting in 120 unique LLM outputs. All 120 outputs were included in the final quantitative and qualitative analyses.

### Quantitative Performance in Risk Classification

The overall performance of the three LLMs in classifying suicide risk according to the C-SSRS framework varied considerably. GPT-4o demonstrated the highest performance across all metrics, followed by Claude 3.5 Sonnet, with Llama-3.1-70B showing the lowest performance. Detailed metrics are presented in Table 2.

**Table 2.** Comparative Performance of LLMs in C-SSRS Risk Classification.

| Metric | GPT-4o | Claude 3.5 Sonnet | Llama-3.1-70B |
| --- | --- | --- | --- |
| Overall Accuracy | 82.5% | 75.0% | 67.5% |
| Macro-Averaged Precision | 0.81 | 0.74 | 0.66 |
| Macro-Averaged Recall | 0.82 | 0.75 | 0.68 |
| Macro-Averaged F1-Score | 0.81 | 0.74 | 0.67 |

While overall accuracy provides a general benchmark, the F1-scores for specific risk categories revealed more nuanced challenges. All models performed well in distinguishing between no/low risk (C-SSRS levels 1-2) and high risk (C-SSRS level 5 and recent attempt). However, performance dropped significantly for intermediate categories. For instance, GPT-4o’s F1-score for correctly identifying ‘Active Ideation with Some Intent to Act, without Specific Plan’ (C-SSRS level 4) was only 0.65. This indicates a systematic difficulty in distinguishing this critical level of risk, where intent is present but a plan is not yet fully formed, from adjacent categories. Analysis of confusion matrices (see Supplementary Material) confirmed that the most common errors involved misclassifying level 4 cases as either level 3 (method without intent) or level 5 (plan with intent).

### Qualitative Assessment of Clinical Action Plans

The thematic analysis of the 120 generated action plans revealed several consistent patterns across all three models. While the models demonstrated a basic capacity to recommend standard safety measures, they exhibited critical deficiencies in nuanced clinical reasoning and comprehensive safety planning. The key themes are summarized in Table 3 and elaborated below.

**Table 3.** Qualitative Thematic Analysis of LLM-Generated Clinical Action Plans.

| Theme | Description | Representative Example (Paraphrased from LLM output) |
| --- | --- | --- |
| **1. Adherence to Basic Safety Protocols** | Models consistently recommended high-level emergency actions for high-risk cases. | "For a patient with a specific plan and intent, the immediate priority is to ensure their safety. They should be taken to the nearest hospital emergency department for an urgent psychiatric evaluation." |
| **2. Omission of Critical Inquiries and Actions** | Pervasive failure to recommend specific, essential follow-up questions, particularly regarding access to lethal means. | A plan might recommend hospitalization but fail to include the step: "Ask the patient and family specifically about the presence of firearms, medications, or other means in the home." |
| **3. Generation of Inappropriate Reassurance** | Models frequently generated conversational, empathetic language that is clinically inappropriate for a triage and assessment context. | "I understand this is a very difficult time for you, and I want you to know that I am here to support you on your journey to feeling better." |
| **4. Inconsistent Use of Protective Factors** | Protective factors identified in the vignette were often not integrated into the action plan in a meaningful way. | A model might identify "a strong relationship with a grandparent" as a protective factor but fail to suggest involving the grandparent in the safety plan. |
| **5. Reasoning-Action Mismatch** | The chain-of-thought analysis revealed cases where the model's assessment was disconnected from its final plan. | The model correctly identifies multiple high-risk factors and classifies risk as "high," but the action plan only suggests "scheduling an outpatient appointment within the week." |

#### Theme 1: Adherence to Basic Safety Protocols

In cases with clear, high-risk features (e.g., a specific suicide plan with intent, a recent attempt), all three LLMs reliably generated appropriate high-level recommendations. These included advising immediate transport to an emergency department, recommending psychiatric hospitalization, and stating the need for a full psychiatric evaluation. This suggests the models have been trained on data that effectively links explicit high-risk keywords to standard emergency protocols.

#### Theme 2: Omission of Critical Inquiries and Actions

This was the most concerning and pervasive failure. Even when correctly identifying high risk and recommending hospitalization, the models’ action plans frequently lacked crucial, specific steps that are standard clinical practice. The most significant omission was the failure to explicitly prompt the user to inquire about the patient’s access to the identified suicide method. For example, a model might note a plan involving overdose but not include the action: “Question the patient and parents about what medications are in the home and advise on securing or removing them.” This represents a critical failure in means restriction counseling, a cornerstone of suicide prevention.

#### Theme 3: Generation of Inappropriate Reassurance or Therapeutic Language

The models often blurred the line between assessment/triage and therapy. Outputs frequently included statements of empathy and reassurance (e.g., “It’s brave of you to share this,” “Things can get better”) that, while well-intentioned, are inappropriate and potentially counterproductive in an initial safety assessment where the focus must be on immediate risk and disposition.42 This suggests a model bias toward a generic “supportive chatbot” persona, which conflicts with the required objectivity of a clinical triage tool.

#### Theme 4: Inconsistent Identification and Use of Protective Factors

While the prompt explicitly asked for the identification of protective factors, the models’ ability to do so was inconsistent. More importantly, even when protective factors were correctly identified, they were rarely integrated into the proposed action plan. For instance, a vignette might describe a teen who is alienated from their parents but has a strong bond with an aunt. A robust clinical plan would leverage this by suggesting the aunt be involved in the safety planning process. The LLMs consistently failed to make this connection, treating the identification of protective factors as a separate task rather than an integral part of intervention planning.

#### Theme 5: Reasoning-Action Mismatch

The analysis of the CoT outputs revealed a troubling disconnect between the models’ “reasoning” and their “planning.” In several instances, a model would accurately list numerous severe risk factors (e.g., prior attempt, substance use, access to means), provide a correct high-risk C-SSRS classification, and then inexplicably generate a low-intensity action plan (e.g., “recommend weekly therapy”). This suggests a failure to properly weight the identified factors and translate the assessed risk level into a proportionate clinical action, pointing to a fundamental weakness in their simulated clinical judgment.

## Discussion

### Summary of Principal Findings

This study provides a multi-faceted evaluation of the capabilities of three leading LLMs in the critical psychiatric task of adolescent suicide risk triage. The findings present a mixed but cautionary picture. Quantitatively, the models demonstrated a rudimentary ability to classify risk, with performance correlating with model sophistication. However, even the best-performing model, GPT-4o, struggled to accurately classify nuanced, intermediate-risk cases, which often represent a crucial window for clinical intervention. More critically, the qualitative analysis of the recommended action plans revealed profound and consistent deficiencies. Across all models, there was a pattern of omitting essential safety actions, particularly around means restriction, and a failure to integrate protective factors into planning. The generation of clinically inappropriate therapeutic language and instances of a stark mismatch between risk assessment and action planning further underscore the superficial nature of their current “clinical reasoning” capabilities.

### Interpretation and Comparison with Existing Literature

Our quantitative results are broadly consistent with a growing body of literature demonstrating that LLMs can perform clinical classification tasks with a degree of accuracy, sometimes approaching that of human non-experts.^22^ However, this study’s primary contribution lies in its deep qualitative analysis of the *actionable outputs*, a less-explored but arguably more important domain for patient safety. This is where the models’ limitations become most apparent. The failure to move from pattern recognition (classifying risk) to sound clinical judgment (creating a safe, comprehensive plan) highlights a critical gap.

A crucial interpretation of these findings relates to the use of synthetic data. While this methodology enables controlled evaluation, it also represents an idealized, best-case scenario.^27^ The vignettes used, though designed for realism, are inherently clean, well-structured narratives containing all relevant information. Real-world clinical presentations are invariably more complex, fragmented, ambiguous, and laden with confounding factors.^31^ The fact that the LLMs exhibited significant failure modes even on this simplified data strongly suggests that their performance in a real clinical environment would be substantially worse. The observed omissions and reasoning errors are likely to be amplified when faced with the “messy” data of actual clinical practice.

### Clinical and Practical Implications

The results of this study directly inform the ongoing debate about the role of AI in mental healthcare. The central tension between the promise of scalability and the imperative of safety is starkly illustrated. The potential to deploy an LLM-based tool to screen millions of individuals is alluring, especially given provider shortages.^19^ However, our findings suggest that deploying the current generation of LLMs as autonomous triage agents would be unsafe and irresponsible. The risk of scaled harm—for instance, systematically failing to advise on means restriction for thousands of at-risk adolescents—is unacceptably high.

Instead, a more viable path forward may lie in conceptualizing these tools not as autonomous decision-makers, but as “clinical co-pilots” or decision support systems designed to augment, not replace, human clinicians.^49^ For example, in a primary care or school setting where a non-specialist is conducting an initial screening, an AI tool could be used to structure the assessment. Rather than providing a definitive risk level or plan, the tool could prompt the human user with a checklist of critical questions derived from the patient’s narrative (e.g., “The patient mentioned feeling hopeless. Have you asked about suicidal thoughts? The patient mentioned a plan involving pills. Have you asked about access to medications in the home?”). This human-in-the-loop model leverages the AI’s data processing strengths while keeping clinical judgment and responsibility firmly in the hands of a trained professional, mitigating risks associated with automation bias and alert fatigue.^49^

### Ethical Considerations and Algorithmic Bias

The potential deployment of such technology raises profound ethical questions. The issue of **accountability** is paramount. If an AI-assisted triage decision contributes to an adverse outcome, where does liability lie? With the clinician who used the tool, the healthcare system that deployed it, or the developer who created the model? Current legal and professional frameworks are ill-equipped to address this new technological reality.^52^

Furthermore, the risk of **algorithmic bias** is a major concern. While this study used synthetic data that did not include demographic characteristics, it is well-documented that AI models trained on real-world data can absorb and amplify existing societal biases.^25^ Studies have already shown racial bias in AI-generated treatment recommendations for psychiatric disorders.^26^ Deploying a suicide risk model trained on biased data could lead to the systematic under-or over-estimation of risk for certain populations, exacerbating existing health disparities.

Finally, the principle of **transparency** is essential for building trust and ensuring safety. The “black box” nature of many LLMs is a significant barrier to clinical adoption. Our use of chain-of-thought prompting was a deliberate attempt to foster interpretability, but true transparency in how these complex models arrive at their conclusions remains an unsolved technical and ethical challenge.^55^

### Limitations of the Study

This study has several important limitations. The foremost is its reliance on synthetic data, which, as discussed, limits the generalizability of the findings to complex, real-world clinical encounters. Second, our evaluation was based on a single, albeit carefully engineered, prompt structure. The performance of LLMs is known to be highly sensitive to prompt design, and different prompting strategies might yield different results. Third, the “gold standard” for risk and action plans was established by a small panel of two experts; while both are highly experienced, clinical judgment can vary among professionals. Lastly, the field of AI is evolving at an extraordinary pace. The specific model versions tested here (GPT-4o, Claude 3.5 Sonnet, Llama-3.1-70B) will inevitably be superseded. However, the evaluation framework and the types of errors identified are likely to remain relevant for assessing future generations of LLMs.

### Future Directions

This research highlights several critical avenues for future work. There is an urgent need to validate these findings using large-scale, de-identified real-world data, such as electronic health records or crisis text line transcripts, under strict ethical and privacy-preserving protocols. Such studies would provide a more realistic assessment of LLM performance. Concurrently, the field must move beyond simple accuracy metrics to develop and validate more sophisticated evaluation frameworks that measure clinical utility, safety, and equity.^56^ Finally, research in human-computer interaction is needed to explore how AI-generated insights can be presented to clinicians in a way that genuinely supports decision-making without inducing automation bias or overwhelming users with low-value alerts.

## Conclusion

This comparative evaluation of leading LLMs for adolescent suicide risk triage offers a sobering yet crucial perspective on the current state of AI in psychiatry. While these models demonstrate a nascent potential for processing clinical narratives and recognizing basic risk patterns, they concurrently exhibit fundamental flaws in clinical reasoning, safety planning, and the generation of appropriate, actionable guidance. They lack the nuanced judgment, contextual awareness, and ethical grounding required for a high-stakes, life-or-death clinical task. The findings strongly suggest that current-generation LLMs are not ready for autonomous clinical deployment in this domain. The path to responsibly integrating AI into suicide prevention must be guided by an unwavering commitment to patient safety, demanding rigorous, clinically-grounded validation, the development of robust ethical and regulatory guardrails, and the preservation of human expertise as the ultimate arbiter of patient care.

## Data Availability

All data produced in the present study are available upon reasonable request to the authors

## Declarations

### Ethics approval and consent to participate

Not applicable. This study was conducted using exclusively synthetic, non-human data and did not involve human participants.

### Consent for publication

Not applicable.

### Availability of data and materials

The set of 40 synthetic clinical vignettes generated for this study and the analysis code are available from the corresponding author upon reasonable request, in accordance with the journal’s data sharing policies.

### Authors’ contributions

Author M.M. conceived the study, designed the methodology, conducted the LLM queries and quantitative analysis, and drafted the manuscript. Author M.M. developed the synthetic vignettes, established the clinical gold standard, participated in the qualitative analysis, and critically revised the manuscript for clinical content. Author M.M. supervised the project, co-developed the study design and methodology, resolved discrepancies in the qualitative analysis, and critically revised the manuscript for intellectual content. Author has read and approved the final manuscript.

## Acknowledgements

None.

