## Supplementary Material for "AI-Powered Triage of Suicidal Ideation in Adolescents: A Comparative Evaluation of Large Language Models Using Synthetic Clinical Vignettes"

### Supplementary Figure S1. Confusion Matrix - GPT-4o

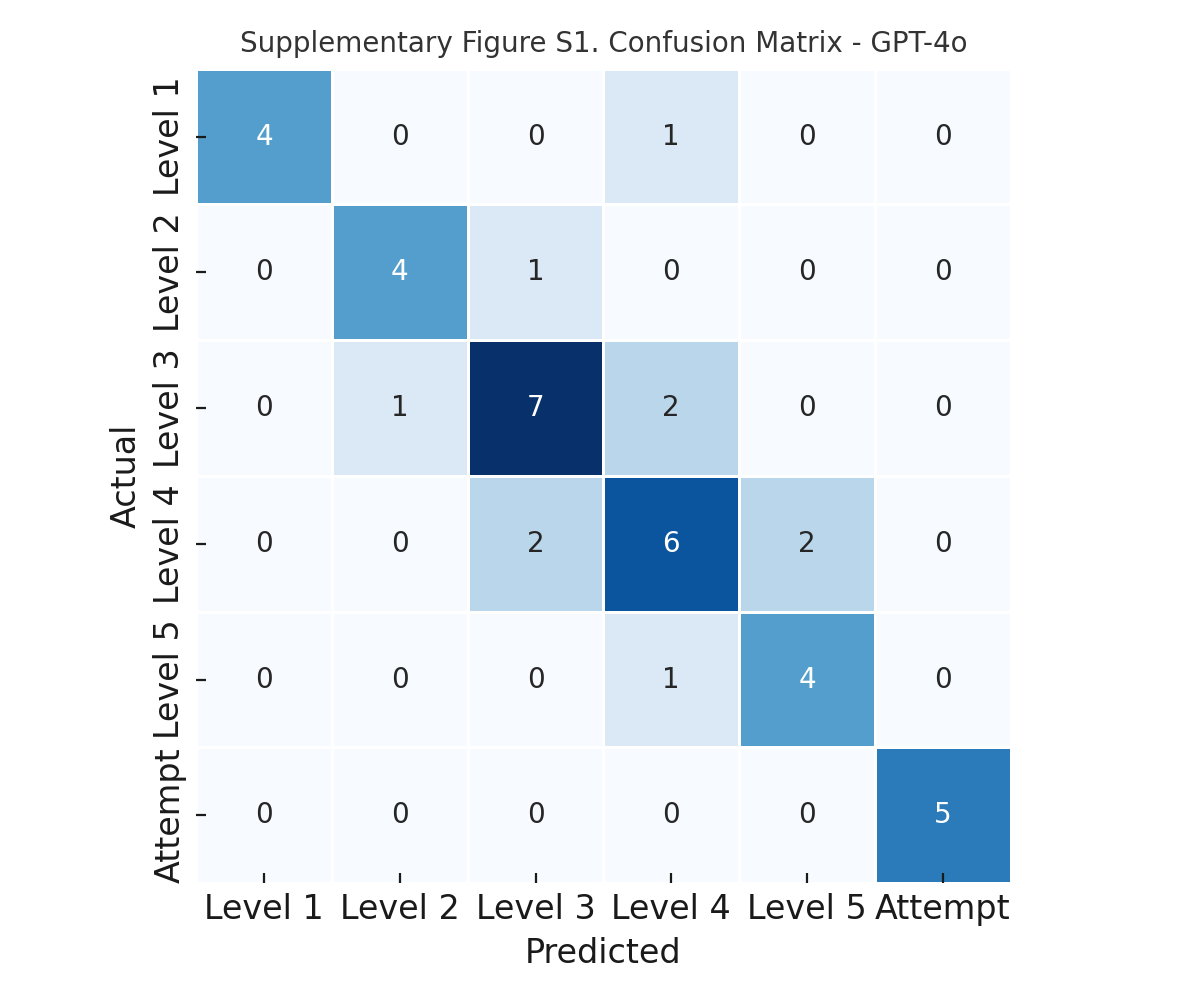

### Supplementary Figure S2. Confusion Matrix - Claude 3.5 Sonnet

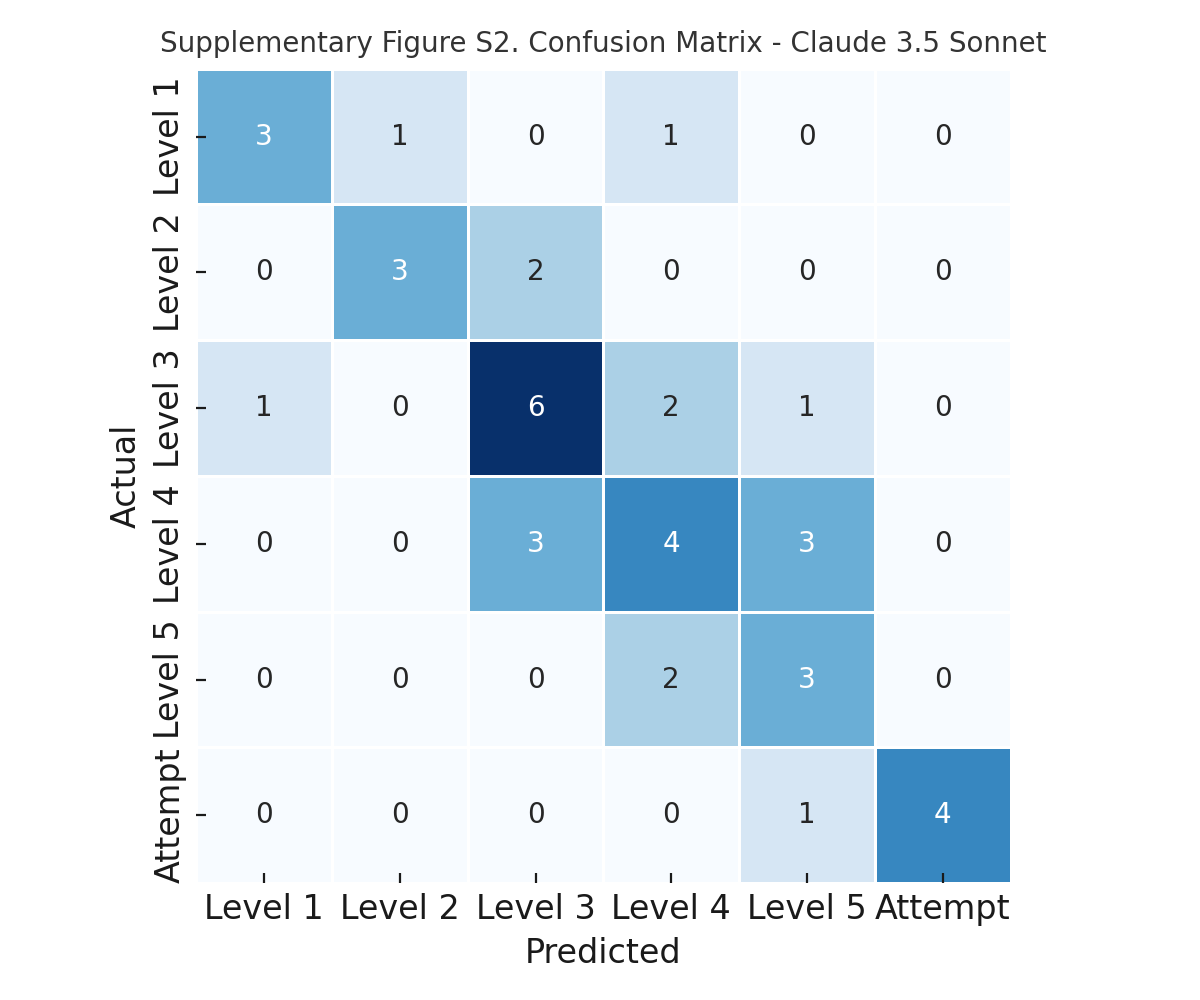

### Supplementary Figure S3. Confusion Matrix - Llama-3.1-70B

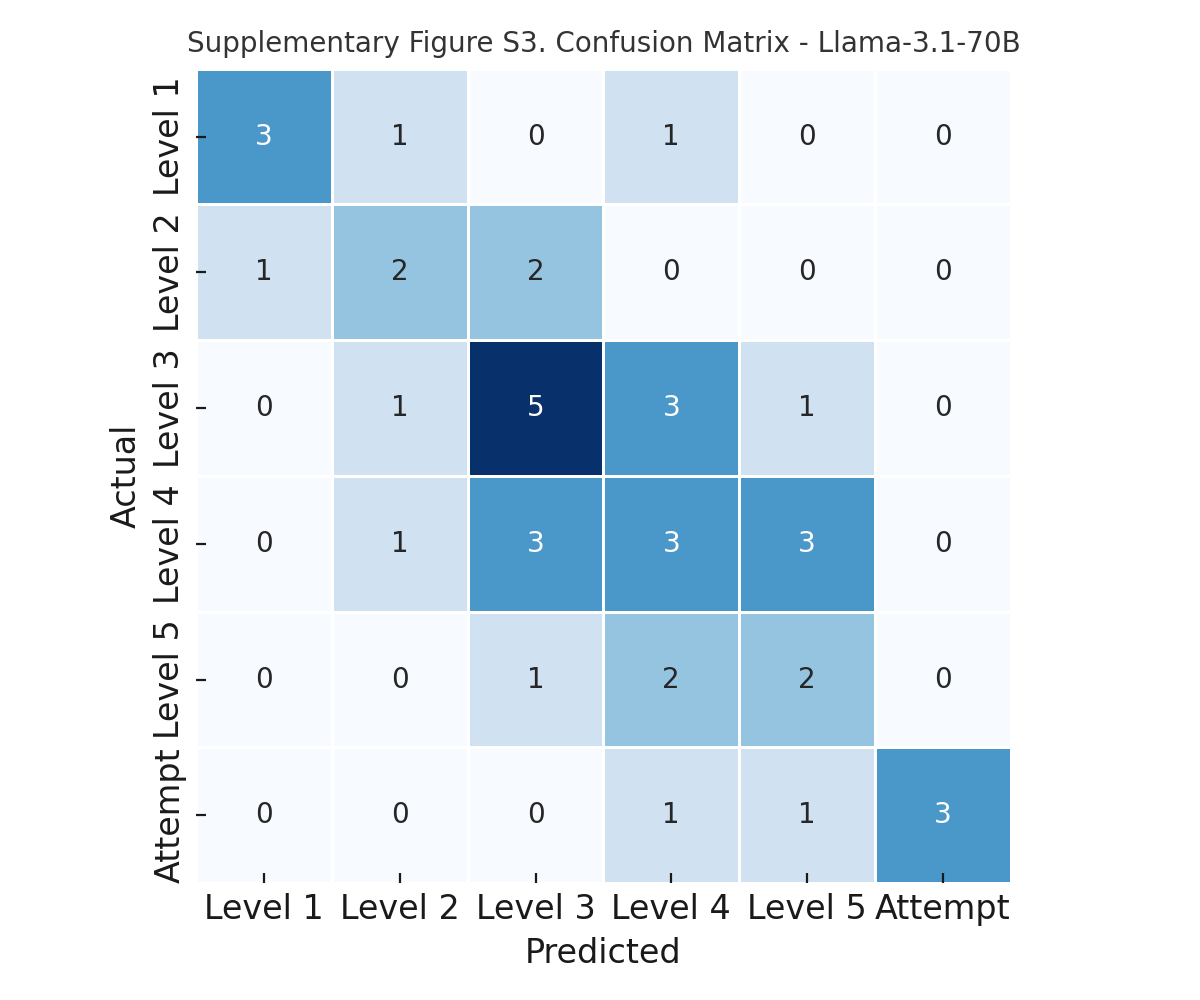
